# Video Directly Observed Therapy for Tuberculosis Treatment in Haitian Prisons: A Pattern of Group Adherence

**DOI:** 10.1101/2023.12.29.23299977

**Authors:** Laura K. Dirks, Patrick M. Bircher, Myrna del Mar González-Montalvo, Alexandra E. Kauffman, Edwin J. Prophete, Margarette R. Bury, Anne C. Spaulding

## Abstract

**Classification:** Research paper

**Purpose:** Haitian provincial prisons historically were strapped to provide directly observed therapy (DOT) for persons with TB (PwTB) due to healthcare understaffing. A non-governmental organization addressed this gap via correctional officer-administered video DOT (VDOT).

**Approach:** A 16-month, quasi-experimental trial of officer-facilitated VDOT started in March 2019 at four prisons. Officers delivered doses directly without video when VDOT was inaccessible. Healthcare staff remotely tracked VDOT adherence asynchronously. Three fully-staffed prisons were controls. Our primary objective was to measure VDOT effectiveness for PwTB who began VDOT within 2 weeks of starting treatment. Our secondary objective was to measure program reach, implementation and maintenance through July 2023.

**Findings:** Reach—55 PwTB on VDOT met study criteria. Effectiveness: median/mean VDOT adherence for 55 individuals enrolled in the pilot were 70.8% and 60.2% respectively. Median/mean total adherence, including doses delivered by officers, were 100% and 93.5%. Implementation: VDOT adherence varied by site but not demographic characteristics; similarity of adherence patterns between subjects within a facility was high. Nursing staff reported that adherence in controls was 100%. Correctional officers reported high comfort with the program technology. Maintenance: Since the pilot, 387 PwTB have received TB medications via VDOT in the Haitian prison system.

**Originality:** VDOT for PwTB in low-resource Haitian prisons enabled close monitoring and follow-up; it could expand treatment options elsewhere. Total adherence neared that in control prisons. VDOT adherence varied by treatment day predominately in a group pattern, reflecting facility-level, rather than individual-level, factors.

## Introduction

Tuberculosis (TB) continues to cause substantial morbidity and mortality in the prisons of low-resource countries worldwide. Inadequate TB care in congregate settings not only threatens the health of persons with TB (PwTB) but also creates a scenario in which TB can spread to others in the facility. While in the past the World Health Organization (WHO) recommended directly observed therapy (DOT) to monitor daily medication adherence, observation alone may not improve treatment completion (Karumbi and Garner, 2015). The importance of supportive healthcare workers in DOT may have been to address selective lapses in drug adherence; incomplete treatment regimens can lead to the development of drug resistance, which VDOT can prevent by incorporating personalized feedback (Ridho et al., 2022). Recent research endorses digital health technologies such as video directly observed therapy (VDOT) as a more convenient and less expensive alternative to DOT (Garfein et al., 2015, Garfein and Doshi, 2019, Garfein et al., 2018, Story et al., 2019) VDOT offers observed therapy without the constraints of a face-to-face encounter between the observer and the PwTB (Vilyte et al., 2023). We sought to demonstrate that VDOT could be implemented to improve treatment outcomes in prisons with insufficient staffing to support nurse-led DOT.

TB transmission is facilitated by crowded, poorly ventilated environments, which commonly characterize prisons. In 2018, the incidence of TB in Haiti was the highest in the Americas, at 176 per 100,000 (WHO, 2019) and its prisons had an even higher prevalence of TB than the general community. Health through Walls (HtW), a non-governmental organization, has partnered with the Haitian National Correctional System since 2001. Its mission is to assist low resource countries in implementing sustainable improvements in the health care services of their prisons (May et al., 2010). In 2017, HtW diagnosed 385 cases of TB in Haiti’s correctional system of 17 prisons. This was not unexpected, given that the correctional facilities in Haiti at the time held roughly 11,000 persons in a system meant for 2,431 (450% capacity) (World Prison Brief, 2023). Prior to 2019, the strategy for treating diagnosed TB cases in Haiti’s incarcerated population was to transfer PwTB for treatment from outlying facilities to the National Penitentiary. Nonetheless, security and resource concerns often prevented transfer, which meant that PwTB remained at correctional facilities that lacked the resources to provide sufficiently observed care and supportive services. While the National Penitentiary in Haiti’s capital, Port-au-Prince, had regular healthcare staffing, many of the provincial prisons did not have nurses on site every day. Nearly 40% (n = 142) of cases remained in resource-poor facilities.

HtW piloted a correctional officer-facilitated VDOT program in select provincial Haitian correctional facilities without the resources to provide supportive DOT via healthcare workers. The program was evaluated for feasibility, reach, efficacy, implementation and maintenance.

## Methods

### Research Setting

Haiti’s office of HtW purchased a portable x-ray machine in 2012, which permitted x-ray screening in multiple prisons across the country. In 2014, a GeneXpert machine for PCR testing of specimens was purchased with funding by USAID and has been in place at the main national penitentiary, the prison in Port-au-Prince, ever since. Systematic, cross-prison testing for TB was conducted using a symptom screen, universal radiography, and sputum testing via GeneXpert for persons with symptoms or radiographic signs of pulmonary TB in 2017. Afterwards, all persons entering prisons in Haiti received the same screening. Persons with HIV infection, sputum positive on GeneXpert and showing rifampin resistance, or both would have specimens sent for culture. For this study, TB cases include both confirmed TB cases (sputum positive on GeneXpert) and probable cases (radiographic or other signs of TB but without demonstration of *M. tuberculosis* either by GeneXpert or culture.)

**Figure 1.**
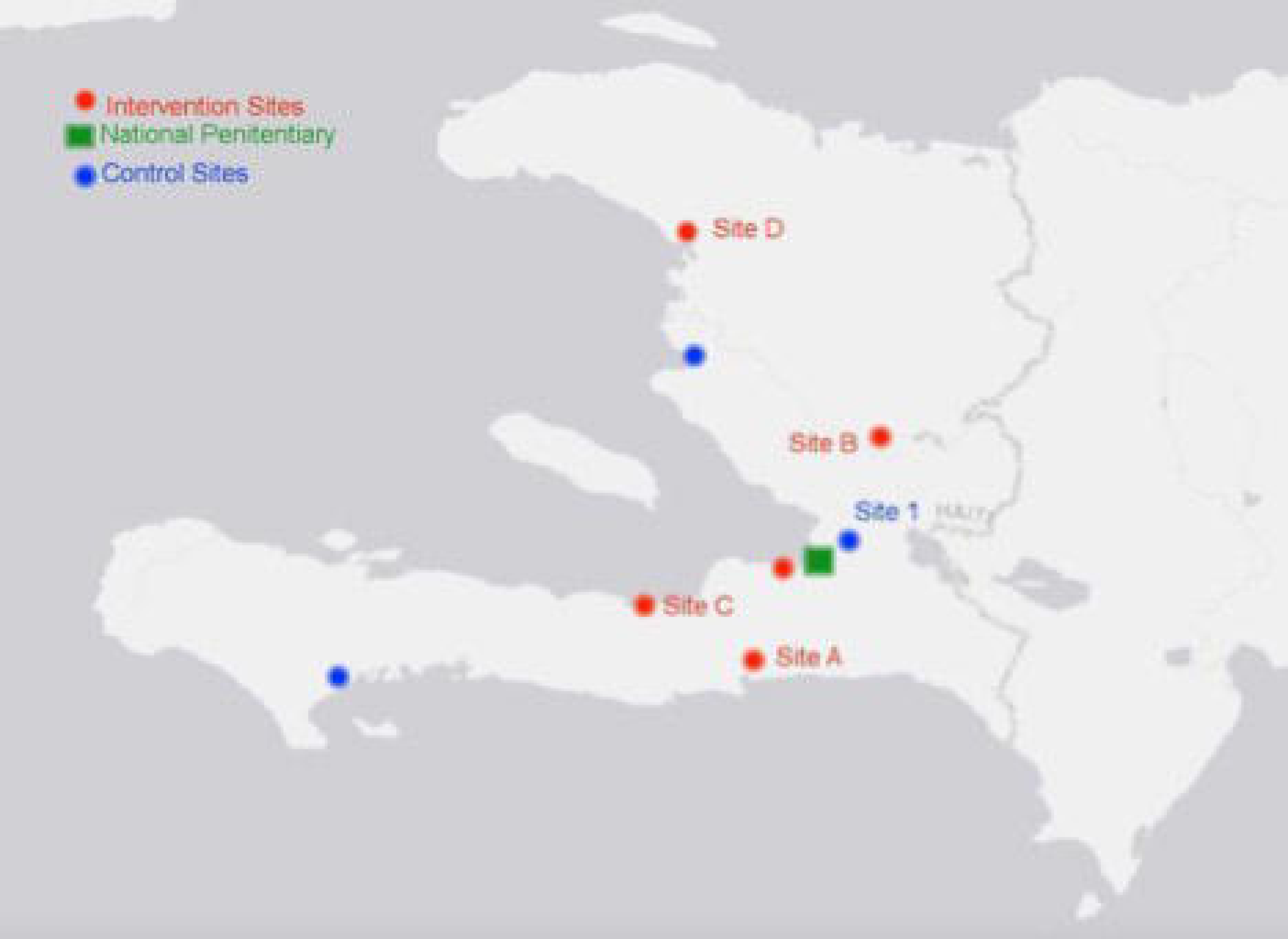

### Study participants

Five prisons were invited to implement the VDOT intervention, and three provincial prisons staffed daily with nurses were chosen as control institutions (Figure 1).

### Intervention

Correctional officers were recruited from the proposed VDOT sites to participate in administering the VDOT program. Benefits of participating included a supplement of up to $50 to their pay each pay period. Each officer who agreed to participate received two days of paid in-person training conducted by the HtW physician. VDOT was delivered using the SureAdhere platform, a smartphone/tablet app developed by researchers at the University of California (Garfein et al., 2018). At the start of the project period in January 2019, the HtW teams began programming the tablets at the study office. One tablet was delivered to each prison. The program assigned a unique PIN for each participant, which automatically linked videos to their adherence records. Officers were trained to escort each participant individually to a private location in the prison to record the PwTB taking their doses, then escort them back to their cells.

We conducted a quasi-experimental trial of adherence to VDOT in prisons where the intervention was the only means of obtaining TB treatment, compared to DOT in control prisons. Control sites consisted of well-staffed correctional facilities able to provide DOT to their populations. In the event that a PwTB on DOT exited the prison system from the National Penitentiary or an intervention prison while on the treatment, they were given the option to transition to VDOT over a mobile phone.

To monitor implementation of VDOT, we will use the RE-AIM implementation framework which evaluates the <u>R</u>each, <u>E</u>ffectiveness, <u>A</u>doption, Implementation and <u>M</u>aintenance of an interventions (Glasgow et al., 1999, Glasgow et al., 2013, Glasgow et al., 2019). Findings on adoption, will be few since the leadership of prisons accepted the invitation for VDOT when no other means of treating TB was available, and staff were paid to participate. See Table 1 for explanation of the RE-AIM measures.

**Table 1.** RE-AIM measure and definitions.

| RE-AIM Dimension | Original RE-AIM Definition | Unit | Definition in this Study | Outcome | Measurement |
| --- | --- | --- | --- | --- | --- |
| Reach | The absolute number, proportion, and representativeness of individuals who are willing to participate in a given initiative, intervention, or program. | Individual | The number and representativeness of VDOT site participants correctional population. | VDOT demographic Data<br>Site Counts | Participant Surveys |
| Effectiveness | The impact of an intervention on important outcomes, including potential negative effects, quality of life, and economic outcomes. | Individual | Adherence and self-reported attitudes about VDOT and technology competency.<br>Quality of life impacts | Proportion of adherence<br>Technology comfort<br>TB stigma scale<br>Depression score | Participant Surveys |
| Adoption | The absolute number, proportion, and representativeness of settings and intervention agents (people who deliver the program) who are willing to initiate a program. | Facility | The number of facilities in which the program is implemented, all other factors being equal. | Proportion of facilities participating in VDOT out of those without DOT | Administrative Records |
| Implementation | At the setting level, implementation refers to the intervention agents' fidelity to the various elements of an intervention's protocol, including consistency of delivery as intended and the time and cost of the intervention. At the individual level, implementation refers to clients' use of the intervention strategies. | Facility | Closeness of protocol implementation to program intention including adaptations and unintended outcomes. Additionally, cost effectiveness of program. | Correctional officer perceptions of the program.<br>Cost of the program<br>Adaptations to protocol and unintended outcomes anecdotally<br>Comparisons between sites | Correctional Officer Surveys<br>HtW costing data<br>Reported adaptations |
| Maintenance | The extent to which a program/policy becomes institutionalized or part of the routine organizational practices and policies. Within the framework, it also applies at the individual level. At the individual level, it has been defined as the long-term effects of a program on outcomes after 6 or more months after the most recent intervention contact. | Individual/<br>Facility | Facility level maintenance including sustainability indicators, and continued stakeholder buy in. | Post-release continuation using VDOT<br>Continued investment from TB Reach<br>Continued involvement of SureAdhere | Adherence data from cell phones<br>Adherence associations<br>Continued support and funding |

### Reach

Reach was defined as the number of persons participating in VDOT, assessed by the number of persons registered on SureAdhere who received their first VDOT dose within 2 weeks of starting TB treatment delivered by officers. Because no other option than VDOT for taking treatment may have been available, we did not compare VDOT versus starts of healthcare worker administered DOT in the same site. We also wanted to understand the characteristics of persons that accessed the intervention compared to the controls. We developed a survey patterned after surveys used in the development of the SureAdhere platform (Garfein et al., 2018) that had questions on demographics, beliefs about TB, and familiarity with cell phone technology. A similar instrument was developed to survey persons residing in control prisons, which did not include questions on familiarity with cell phone technology. Study staff collecting human subjects data were Haitian Creole speaking and trained in the CITI research ethics program (Fort Lauderdale, FL). Persons who were at the intervention and control prisons currently receiving TB treatment were eligible to participate in the survey, and a convenience sample was recruited at each, dependent on who was receiving treatment on the days the surveyors visited. Written informed consent to participate in a survey, a focus group, or both was solicited before collecting any human subjects data. Those who chose not to participate in the evaluation were free to continue their current treatment with their available care, either VDOT at an intervention prison, or daily DOT at a control prison. After survey completion, participants in the residents’ survey were compensated in the form of a hygiene package.

### Effectiveness

The primary aim of this study was to evaluate the effectiveness of VDOT within correctional populations of Haiti. For this study, effectiveness is defined as taking a TB regimen via VDOT, as prescribed. Adherence is defined as the percentage of planned doses of the TB regimen taken at the time planned. The effectiveness of VDOT was assessed by determining the mean percentage of doses by VDOT and the VDOT adherence fraction (VAF) by each intervention site. The total adherence fraction (TAF) for each intervention site was also calculated to assess the ability to continue providing treatment in the event of technology failure. We wanted to understand whether adherence to TB therapy was significantly associated with demographic characteristics of enrollees. We sought to understand if adherence to TB therapy was correlated with the enrollees’ perceived TB stigma and mental health. The survey provided a measurement of perceived TB stigma adapted from an existing instrument used to measure TB related stigma in Haitians living in the Léogâne Commune of Haiti (Coreil et al., 2010a, Coreil et al., 2010b). TB stigma was measured using a scale of 15 items reflecting five stigma components: internal shame, disease disclosure, external concerns, family reputation, and assumption of other illnesses. (Table 2; details are presented elsewhere (See preprint in acknowledgements). In addition to stigma, we also evaluated the possible presence of depression of participants using a validated screening instrument for depression developed by Partners in Health in Haitian Creole (Rasmussen et al., 2015). Based on the initial study of the Zanmi Lasante Depression Symptom Inventory (ZLDSI), a score between 12 and 14 is parsimonious to depression measures in U.S. scales (Rasmussen et al., 2015). Details about the use of the ZLDSI in this study are provided elsewhere (See preprint in acknowledgements). Further psychiatric work-up was unavailable. We sought to gain participant feedback on use of VDOT by conducting focus groups of 6-12 consenting participants in both VDOT and control prisons. Groups were facilitated by Creole speaking facilitators, recorded, and recordings were transcribed. In the private, common areas where groups assembled, background noise in the prisons precluded meaningful interactive discussion among the participants and no further groups were conducted after the first three were attempted. No coding of transcripts was undertaken.

**Table 2:** Stigma Measures. (Source: Adopted from Coriel *et al.)*

|  |
| --- |
| Internal Perceptions and Emotions |
| 1. Do you think less of yourself? For example, has it reduced your pride or self-respect? |
| 2. Have you ever been made to feel shamed or embarrassed because you have TB? |
| Disclosure |
| 1. If possible, would you prefer to keep people from knowing that you have you have TB or have been assigned a cell for people with TB? |
| 2. Have you discussed or do you plan to discuss with your family that you have TB? |
| 3. Have you discussed or do you plan to discuss with other people detained here that you have TB? |
| 4. Have you discussed or do you plan to discuss with guards that you have TB? |
| External Perceptions |
| 1. Do you think that other persons detained, others in the prison, or others in your community have less respect for you? |
| 2. Do you think that other persons detained, others in the prison, or others in your community have less respect for your family? |
| 3. Do you think people are likely to think you have other health problems (even if you don't)? |
| 4. Are you worried that people will assume you have HIV? |
| External Actions |
| 1. Do you feel others have avoided or might avoid you? |
| 2. Do you think some people refuse to visit your home because of TB even after you have been treated? |
| 3. Have you been asked to stay away from work or social groups? |
| 4. Do you think you might decide on your own to stay away from work or social groups? |
| Courtesy stigma |
| 1. Do you think contact with you might have any bad effects on others around you, even after you have been treated? |

### Adoption

Not addressed. Adoption would be reflected by the proportion and representativeness of prisons and workforce choosing to participate in the VDOT program. A study design where investigators paid officers to participate precluded measurement of whether the prison staff embraced the VDOT technology based on its intrinsic merits.

### Implementation

At the facility-level, implementation was measured by the percentage of days all PwTB received their doses using the same modality. On an individual level, we noted the number of participants who did not finish the protocol. We also sought to evaluate implementation using quantitative and qualitative data collected from correctional officers. Correctional officers were compensated $5 if they engaged with the evaluators. Custody views were measured through specific questions on a survey instrument for correctional officers. We also sought officer feedback on use of VDOT by conducting focus groups of consenting staff. One focus group of 3 officers was assembled in a VDOT prison. The group was facilitated by a Creole speaking facilitator and recorded. The recording was transcribed. Due to the inability to assemble additional focus group because of background noise of the prisons (see above), further analysis of the transcript was not performed.

Finally, we measured challenges with implementing the VDOT protocol and unintended outcomes. Cost of VDOT in the Haitian prisons was also used as a measure of implementation.

### Maintenance

On an individual level, we measured the continuation in using the VDOT app by released PwTBs after they returned to the community and whether they completed treatment. On an institutional level, the number of PwTBs who continued to be started on the SureAdhere VDOT platform after the pilot study until July 2023 was tabulated. Because no other option for taking treatment may have been available, we did not compare VDOT versus DOT starts at the same site in the maintenance phase.

### Statistical collection and Analysis

Inclusion criteria for analysis required that beginning at each prison’s start date for implementation of VDOT, PwTBs must utilize the VDOT within 2 weeks of starting any DOT. Data from the instruments were transferred to a REDCap (Research Electronic Data Capture) tool hosted at Emory University (Harris et al., 2019, Harris et al., 2009).

The VAF was defined and calculated as the number of VDOT doses taken, divided by the expected number of treatment days. The TAF was defined and calculated as the number of total doses (DOT and VDOT) taken, divided by the expected number of treatment days. One-way ANOVA was used to determine whether there were significant differences in VAF and TAF between intervention sites. Lastly, the mean percentage of doses by VDOT was calculated by dividing the total number of VDOT doses given at each site by the total number of doses given.

Associations between adherence and demographic variables were assessed using a simple linear regression. The combined data was analyzed using R. Students T-tests were used to determine differences in sociodemographic characteristics for VDOT and DOT sites. A T-test of average stigma scores for those enrolled in the VDOT study compared to DOT participants was performed to investigate if there was a significant difference in TB stigma between the groups.

### Ethical Considerations

Study protocol was reviewed and approved by the Emory University Institutional Review Board (IRB). Locally, the Haitian National Bioethics Committee also approved the study protocol. Additionally, The National TB Program in Haiti (PNLT) approved the study and voiced support of the protocol in the context of Haitian Prisons. All documents used to collect participant information--consent forms, surveys and focus group scripts--were translated into Haitian Creole, and back translated; they were read aloud to ensure full understanding and informed consent.

## Results

### Reach

Five prisons were invited to start the intervention. Officers volunteered to participate in the VDOT program in 4 of the 5. From March 2019 – July 2020, 73 imprisoned persons enrolled on the SureAdhere platform. There was one loss to follow up and one death before VDOT was started. Fifty-five persons receiving VDOT in these 4 prisons met our inclusion criteria for analysis. In comparison, a convenience sample of an estimated 102 persons started on DOT in the control prisons.

Participation was high in the VDOT program, where there was no alternative means of treatment. Six residents finished their course after exiting the prison. The enrolled population had a mean age of 31 years at their start date [range 18 – 59 years] and were 100% male.

Twenty-eight (50.9%) participants from the 4 VDOT sites completed surveys. Ten participants from one DOT control site (see map: Site 1) completed surveys. Political unrest and gang warfare during the study period, followed by the COVID-19 pandemic, prevented evaluation staff from reaching sites and making planned follow-up visits. The study staff offered and conducted the survey with only a limited number of individuals.

Of those surveyed in VDOT prisons, all (28/28) had no concerns regarding participation in the VDOT program. Participants had high knowledge of TB; all 28 identified that TB is curable, and most (23/28) correctly identified at least one symptom. Most (26/28) also identified at least one treatment method for TB. The majority of the group (20/28) believed TB is a ‘very serious’ disease, and most (22/28) believed TB is a ‘very serious’ problem in Haiti.

### Effectiveness

The median total adherence fraction (TAF) for both intervention and DOT control sites was 100%. Among the intervention prisons, sites A, C, and D had the highest median TAF of 100% (SD: 0, 16.9, and 17.8, respectively), followed by site B with median (SD) TAF of 98.3% (18.2). There was no significant difference in TAF by intervention site. Site A had the highest mean TAF of 100%, followed by sites D, C, and B of 94.0%, 92.9%, and 89.7%, respectively.

The median VDOT adherence fraction (VAF) across all 4 intervention sites was 70.8%. Analysis of adherence and demographic variables found no significant associations between age, education level, or TB knowledge with VDOT adherence. While there was no significant difference in TAF by intervention site, VAF varied significantly by site. Site B had the highest median (SD) VAF of 87.7% (24.4), followed by sites D, A, and C, with median (SD) VAF of 77.8% (30.2), 65.1% (19.8), and 18.3% (37.7%), respectively (Table 1). Site B had the highest mean VAF of 76.3%, followed by sites A, D, and C, with mean VAF of 72.2%, 66.4%, and 39.8%, respectively (Table 3).

**Table 3:** TB regimen adherence by prison site, Haiti, 2019-2020.

|  | Total | Site A | Site B | Site C | Site D | P-value<sup>1</sup> |
| --- | --- | --- | --- | --- | --- | --- |
| No. VDOT PwTBs | 55 | 9 | 13 | 18 | 15 | — |
| Total Adherence,<br>Median % (SD) | 100<br>(16.1) | 100 (0.0) | 98.3 (18.2) | 100 (16.9) | 100 (17.8) | 0.568 |
| Total Adherence,<br>Mean % | 93.5 | 100 | 89.7 | 92.9 | 94.0 | — |
| VDOT Adherence,<br>Median % (SD) | 70.8<br>(33.6) | 65.1 (19.8) | 87.7 (24.4) | 18.3 (37.7) | 77.8 (30.2) | 0.004 |
| VDOT Adherence,<br>Mean % | 60.2 | 72.2 | 76.3 | 39.8 | 66.4 | — |
| No. participated in<br>baseline survey | 28 | 7 | 7 | 12 | 2 | — |

<sup>1</sup>one-way ANOVA

Across the participating intervention sites A, B, C, and D, mean percentage of doses by VDOT was 66.9%, 80.7%, 54.8%, and 28.1%, respectively (See Figures 2a-2d).

**Figure 2a-2d.**
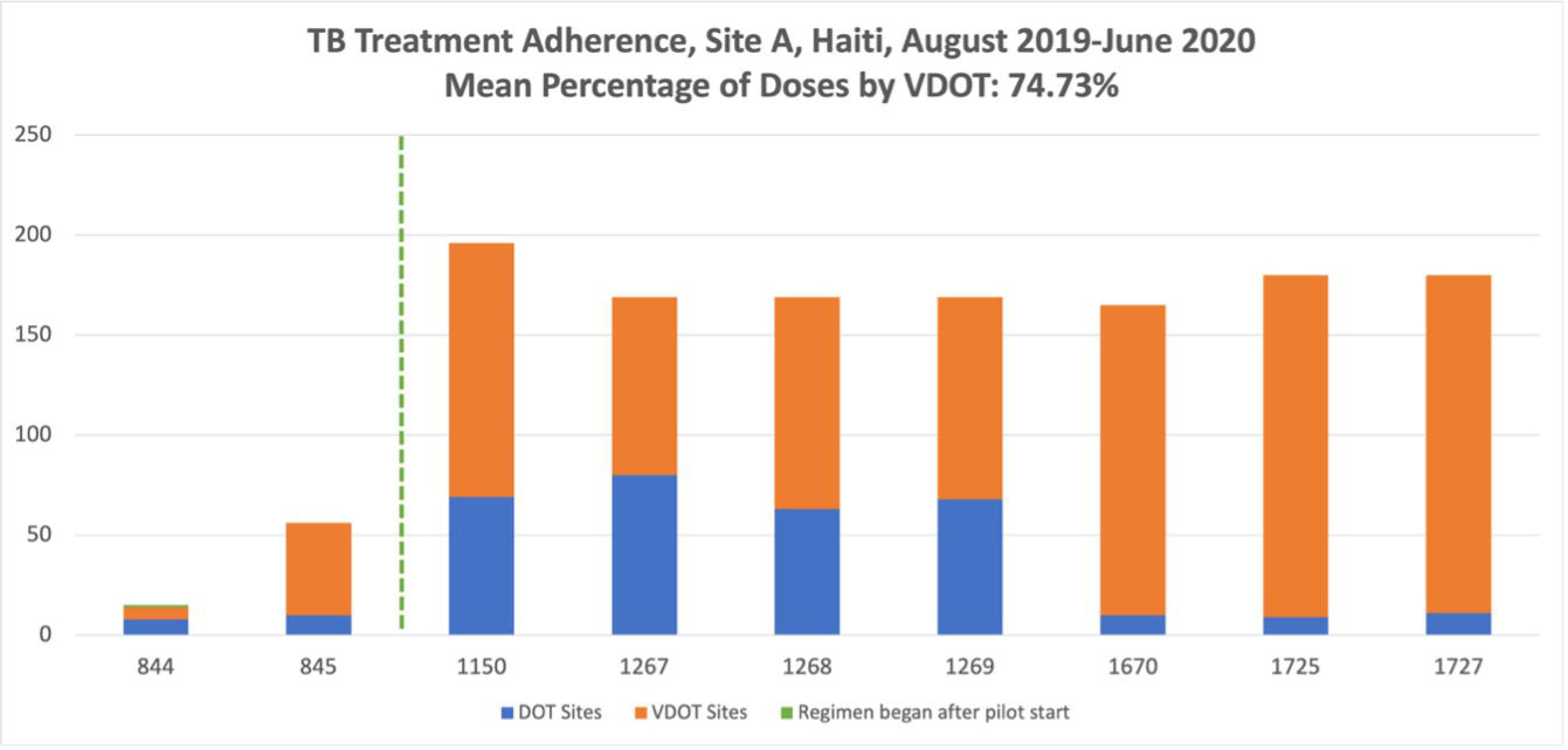
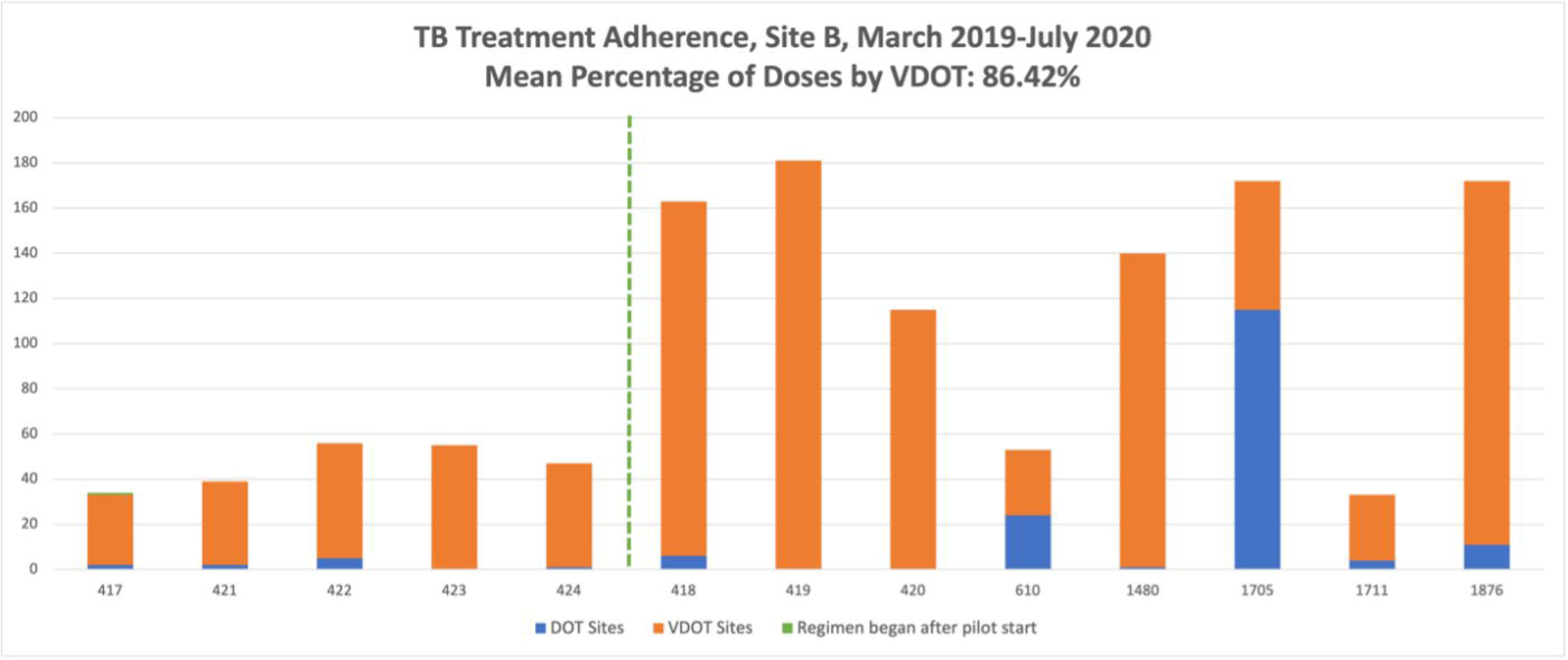
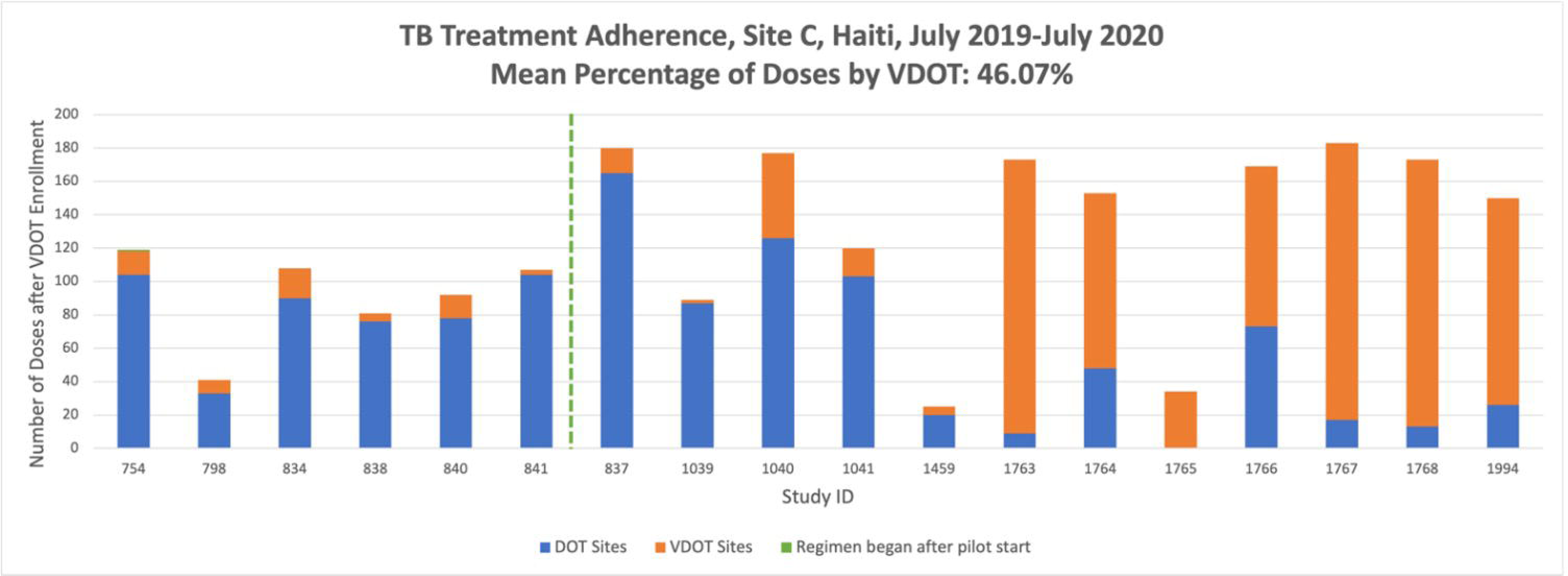
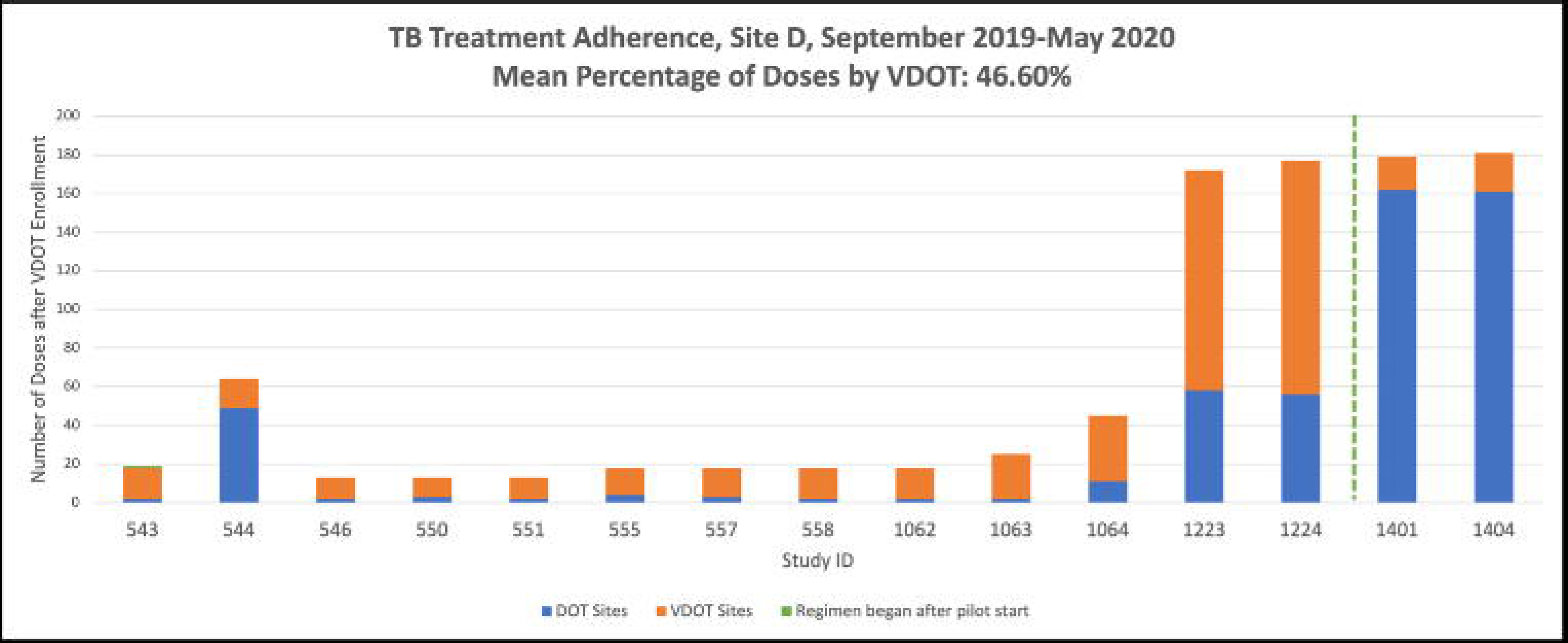

The mean (SD) TB stigma score for VDOT was 39.4% (20.6), which did not significantly differ from the average DOT stigma score of 34.4% (14.5). Among the 16 participants who completed the stigma survey, there was a significant positive linear association found between TB Stigma Score and VDOT adherence.

Average depression scale values for VDOT participants (16.3) and DOT participants (15.7) were not significantly different. Additional evaluation of the depression scale is summarized elsewhere (See preprint in acknowledgements). Follow-up survey results found that 5/6 participants “did not mind the VDOT process”; one participant felt it made them feel “like I am not trustworthy”.

### Adoption

Adoption would be reflected by the proportion and representativeness of prisons and workforce choosing to participate in the VDOT program and could not be formally measured due to the study design.. Regarding representativeness of the prisons and their participants, the VDOT population did not differ significantly from the DOT population for measures of age, years of school, TB knowledge, or Stigma Score (Table 4), but the sample was too small to draw meaningful conclusions.

**Table 4.** Demographic data of VDOT and control prisons, Haiti, 2019-2020.

|  | VDOT Prisons (Sites A-D) | Control Site (Site 1) | P-value<sup>1</sup> |
| --- | --- | --- | --- |
| Age, Mean (SD) | 31.1 (8.75) | 30.2 (6.44) | 0.735 |
| Years of School, Mean (SD) | 3.91 (3.91) | 6.8 (4.52) | 0.099 |
| TB Knowledge Proficiency, % of survey takers | 67.9% | 40% | 0.156 |
| Comfort with Cell Phone Use Score<sup>2</sup>, Mean (SD) | 5.4 (3.24) | — | — |
| TB Stigma Score<sup>3</sup>, Mean (SD) | 39.44 (20.63) | 34.44 (14.49) | 0.476 |

<sup>1</sup>Two-sample T-test.
<sup>2</sup>Cell phone use comfort score ranged from 1 (very uncomfortable) - 10 (very comfortable).
<sup>3</sup>TB Stigma Score was scored from 0 (no stigma) - 100 (high stigma)

### Implementation

An attempt to gather qualitative information on the workforce was not sufficiently large to reach conclusions. Correctional officer recruitment and participation was a barrier to enrolling at one of the sites. However, positive feedback was received from recruited officers. All (3/3) correctional officers participating in a focus group at a VDOT site “completely agreed,” they would participate in increasing accessibility for treating a disease, “even if they doubted the efficiency in correctional institutions.” The group also completely agreed that “My principles are to make treatment accessible for inmates,” but two strongly disagreed that it is morally unacceptable to block medical treatment for inmates. Finally, all three correctional officers strongly agree that they feel proud to be increasing the availability of tools to promote adherence to medical treatment.

Correctional officers reported intermittent difficulty connecting with the internet and occasional tablet malfunction, which would lead to a pattern of all or no adherence to VDOT. In the intervention prisons, VDOT vs. officer-administered DOT adherence on a given day was largely similar for all participants, based on internet connectivity. Analysis of the intervention prisons showed that regardless of the therapy modality, on 75.7% of days (weighted average across the 4 intervention sites), all PwTB on treatment received their doses using the same modality. By site, PwTB received their doses by the same modality 85.1%, 73.7%, 67.0%, and 82.4% of days at sites A – D, respectively (See Figures 3a-3d for snapshots of group adherence patterns). The Appendix contains complete medication administration records for all subjects of this study.

**Figure 3a-3d.**
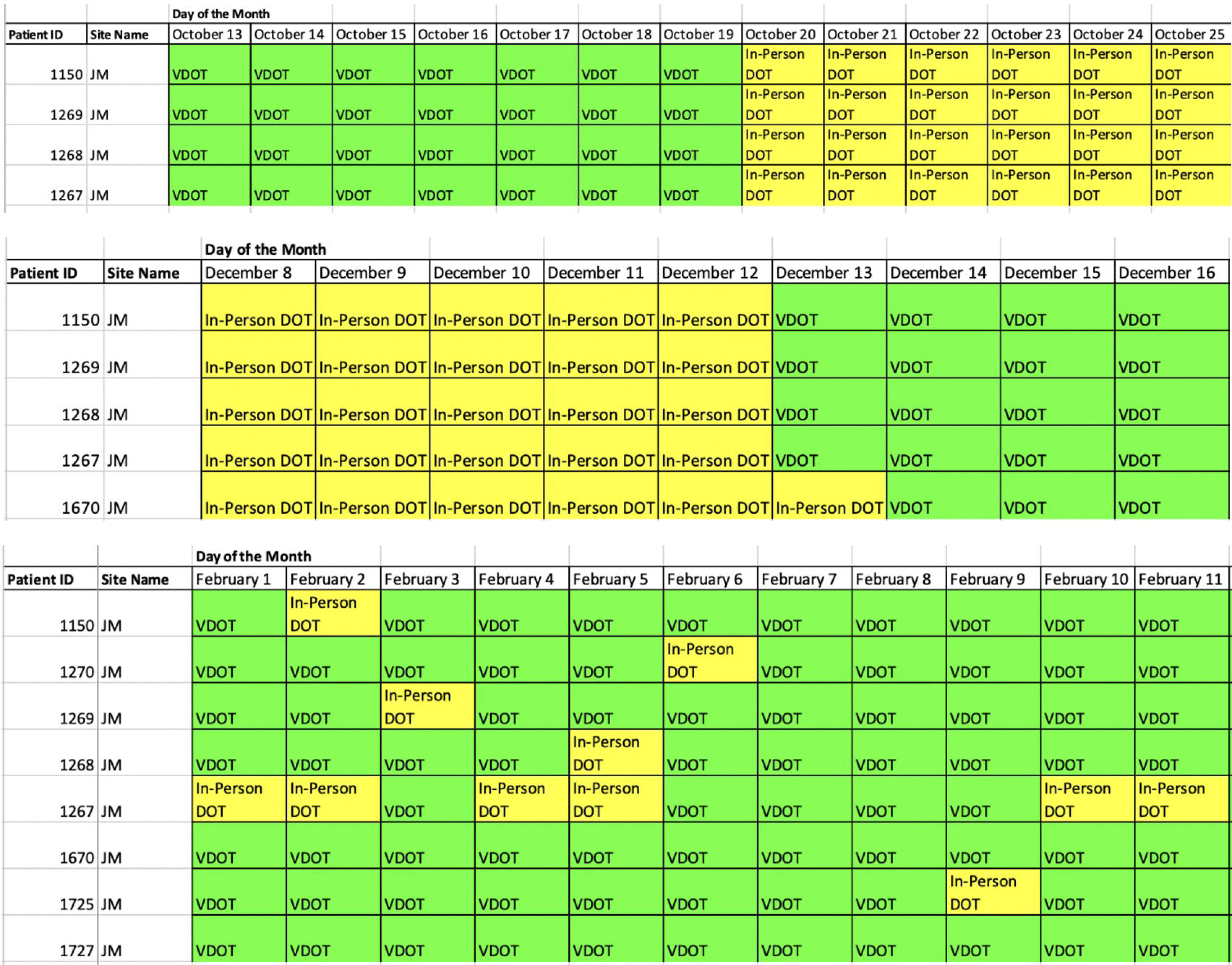
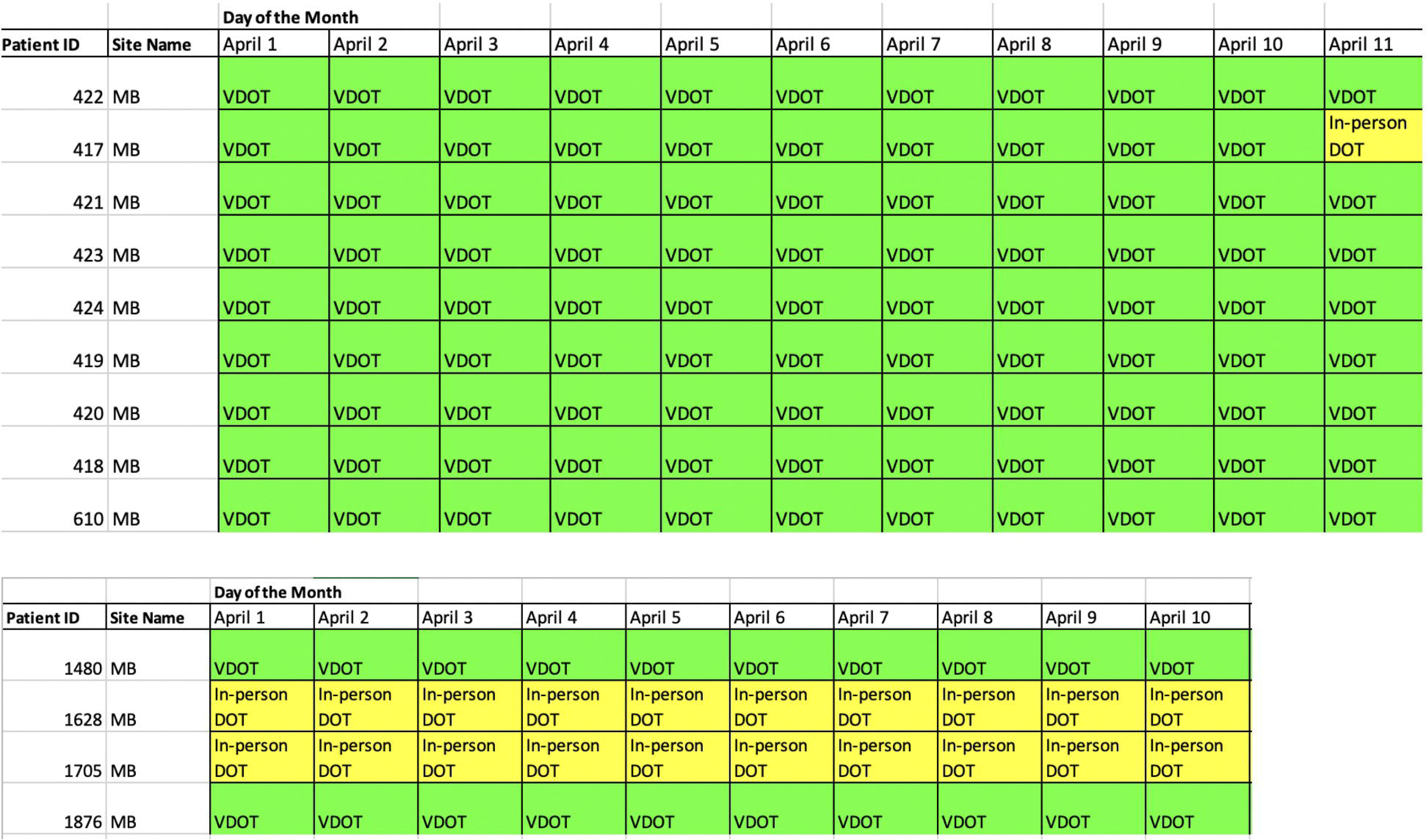
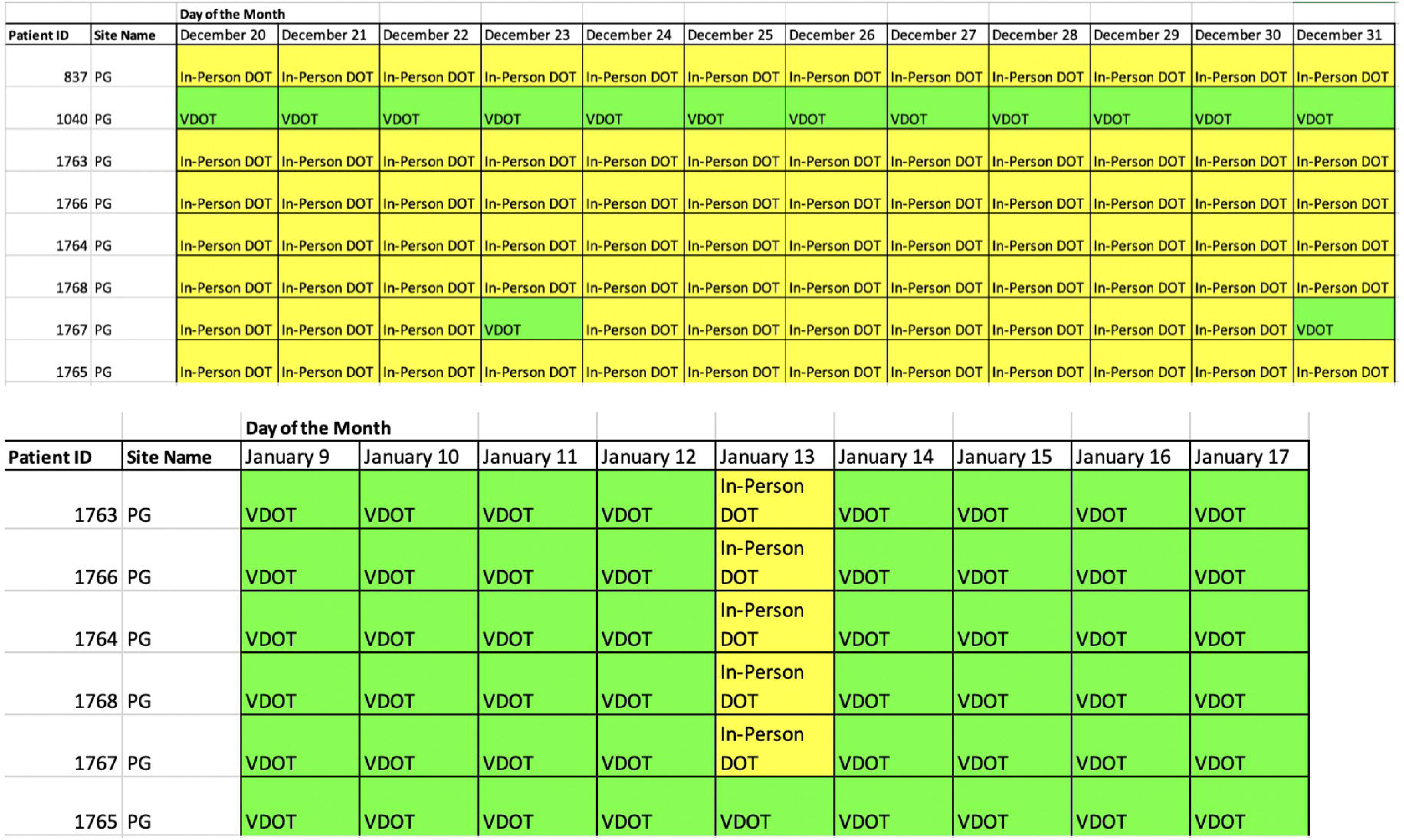
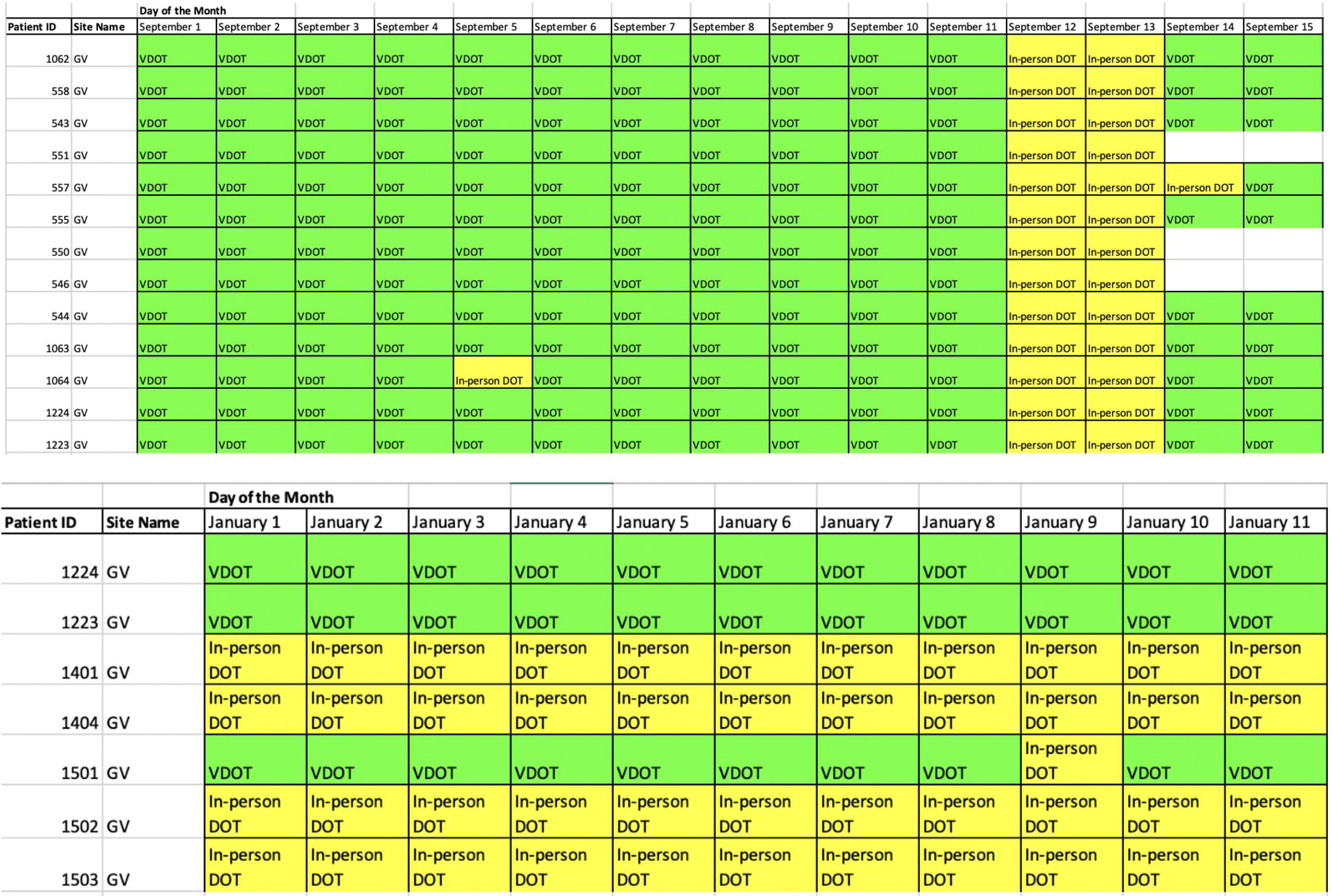

Correctional officers reported three main challenges in implementing VDOT: maintaining a continuous network connection; civil unrest causing complete lockdown or limiting road access; and the tablets having malfunctions, which caused a delay in video uploads or missing videos. During implementation, two sites struggled with tablet malfunctions, which produced an error message when launching the application used for VDOT. These tablets were replaced with upgraded devices that resolved the problem. Other sites reported occasional network and connectivity issues. In the event of network issues, videos were stored on the devices and uploaded once reconnected.

One site reported a secondary use for the camera of the provided tablet, to create a photo registry of those incarcerated at their site. The VDOT program, minus the evaluation, costs significantly less than its DOT counterpart. Initial equipment was around $200 per tablet. Monthly costs included internet access of $8 - $10 a month depending on carrier, and correctional officer compensations. Correctional officers are compensated through a percent increase in their monthly salary, which varied but did not exceed $50 per officer. Adding full-time nurses at the five outlying prisons would cost around $6,000.00 annually, or an average of $500 monthly per site. Based on monthly estimations, the VDOT first-year program costs were $8,080, compared to $30,000, had full-time nurses been available and hired.

### Maintenance

#### By individual

Once participants completed their treatment, they no longer needed intervention. Of those given a cell smartphone to continue VDOT after exiting correctional facilities, 2 admitted to selling their phones due to financial strain, and 1 individual lost their phone. Many reported network issues and other technical difficulties. Six individuals who maintained possession of their phones were included in this evaluation. Four individuals exiting site C completed treatment, and 2 individuals exiting the National Penitentiary were still using VDOT at the time of the evaluation. The median TAF for this group was 57.6%, and median VAF was 47.3%. Medians for the National Penitentiary (68.9% TAF and 52.5% VAF) were higher than those leaving site C (53.5% TAF and 39% VAF). Despite low recorded adherence, all 4 participants from site C completed treatment.

#### By program

After the pilot phase of the study, maintenance was measured for the following 3 years. Over this time period, HtW continued the SureAdhere program for PwTB (Table 5).

**Table 5.**
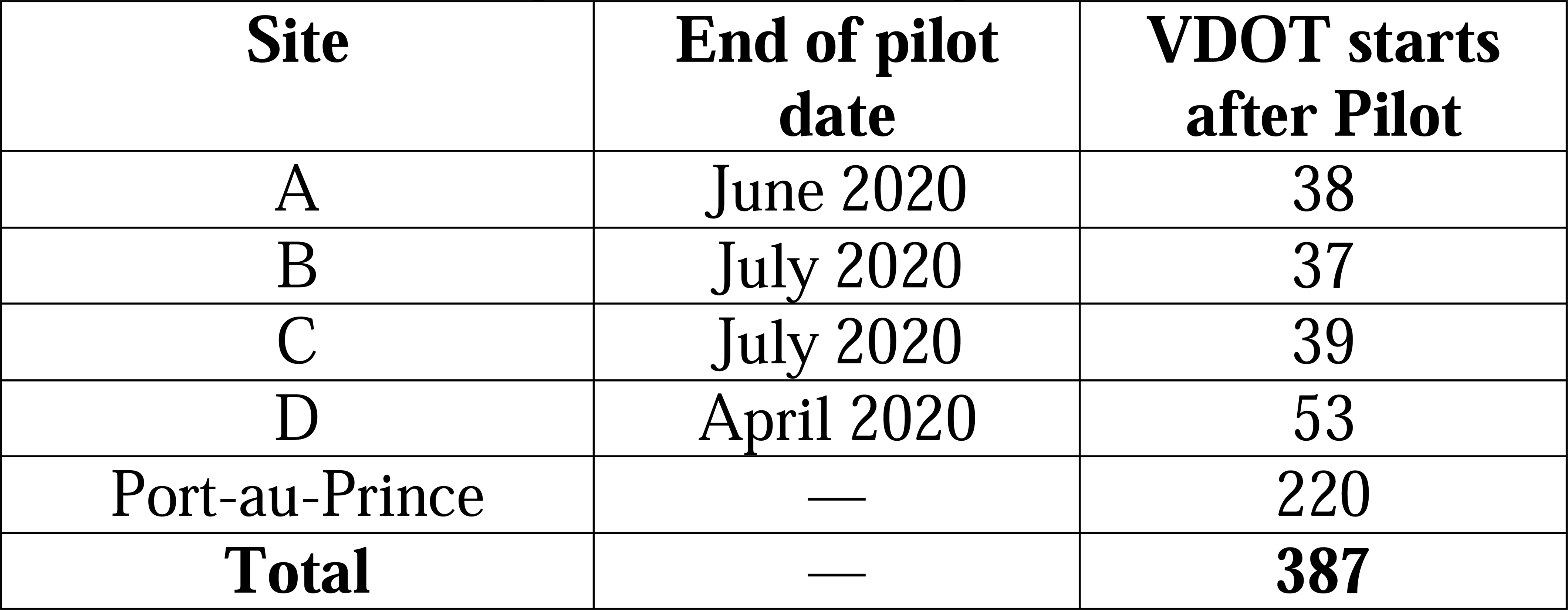
VDOT Starts in Study Prisons of Haiti, from after Completion of Pilot, May 2020 until July 2023.

Access to remote prisons has not improved in the subsequent years, which have been marked by continued gang warfare and the Haitian president’s assassination in 2021.Transportation for healthcare workers to both provincial prisons and the National Penitentiary worsened after the pilot program; the continued uptake of VDOT provided a means to continue treating PwTB rather than abandoning treatment.

## Discussion

The median VAF, 70.8%, was lower than previous studies conducted in higher resource settings (Garfein et al., 2015). When factoring in doses marked as DOT by correctional officers, the TAF increased to a median of 100%. Our analysis shows that adherence in the prisons followed a group pattern—on 41% of days, all residents received either VDOT or officer-administered medications.

A previous publication reported the results of this and several other VDOT studies in various settings in low resource countries (de Groot et al., 2022). The study described in this report was the sole project in a prison. While this prison program had the lowest adherence rate, the combination of VDOT punctuated by officer delivered medications without VDOT neared that of control facility. VDOT is a tool that has been proven effective in increasing treatment completion (Karumbi and Garner, 2015). Linkage with the central office of HtW could provide support for PwTB that would have not been available in the absence of VDOT, if officers managed the entire treatment program alone, without VDOT technology. The future goal for VDOT in the prisons of a low resource country may be as a check for “most” doses, with an understanding that challenges with maintaining internet connectivity may preclude 100% of doses being monitored. Even with connectivity issues, VDOT provides the means for PwTB to report side effects and other challenges with taking TB regimens. The fact that HtW has used VDOT in prisons for the three years following the study is testimony to its usefulness.

Sites A and C had the lowest median VAF of 65.1% and 18.3%, respectively. Still, both sites had a median TAF of 100%. The VDOT population reports no difference in TB stigma, age, education, or TB knowledge than the DOT population. Disclosure, which is a measure of willingness to discuss TB diagnosis, is highly related to internal perceptions (Coreil et al., 2010a, Coreil et al., 2010b). The selling of phones indicates that the population may need further financial incentives to succeed. Despite these challenges, and low adherence, those who continued using smartphone VDOT did complete treatment.

Limitations include the measurement of reach. We could measure how many individuals enrolled in the program but not how VDOT increased treatment completion over the usual TB program. The lack of alternatives to VDOT in the intervention prisons meant there was no choice for targeted PwTB. The inability to measure adoption was inherent in the study design.

The success of correctional officer facilitated VDOT has the potential to close the gap in TB care experienced by low resource correctional facilities. Long-term maintenance of the program will depend on correctional officer buy in. The evaluation of this program can be used to inform future scale-up activities in Haiti. The pilot study and program follow up were designed to demonstrate the value of implementing VDOT for current and formerly incarcerated individuals in low resource settings.

## Recommendations

VDOT could be implemented in other countries where staffing of health care services in prisons is insufficient. VDOT is a promising method to deliver TB care for those transitioning out of prison. The TB stigma scale used in this study indicate there is a high level of stigma in both VDOT and DOT populations.

## Conclusions

VDOT facilitated by correctional officers can provide TB treatment in low resource carceral facilities. It extended the reach of HtW’s TB program into rural prison settings. Not only did it facilitate observed therapy, but it also enables continuous monitoring of PwTB and follow-up on treatment that historically was extremely challenging for these prison sites. Adherence to VDOT followed a group rather than an individual pattern, implying that its use versus non-use depended on facility level factors rather than individual preference of PwTB. Implementing VDOT for TB therapy could expand treatment in correctional facilities in Haiti and be used in prisons in other low resource areas facing similar challenges.

## Data Availability

All data produced in the present study are available upon reasonable request to the authors.

## Acknowledgements

We wish to thank Kelly Collins PhD, Shanaika Grandoit MPH. This work was supported by the Center for AIDS Research at Emory University (P30AI050409). We also appreciate the support of the TB Research Advancement Center (TRAC, P30AI168386) of Emory/Georgia.

